# Indocyanine Green Fluorescence-Guided Sentinel Lymph Node Biopsy in Breast Cancer Using the MARS Near-Infrared Imaging System: A Prospective Single-Center Feasibility Study

**DOI:** 10.64898/2026.06.30.26356987

**Authors:** F. Kurdi, Y. Kurdi, M. Kurdi, T. N. Pisareva, N. S. Sukortseva, A. A. Shiryaev, A. L. Istranov, I. V. Reshetov

## Abstract

**Background:** Sentinel lymph node biopsy (SLNB) is an essential component of axillary staging in breast cancer. Fluorescence-guided mapping with indocyanine green (ICG) enables real-time visualization of lymphatic drainage; however, the parameters of fluorescence-signal recording require standardization.

**Objective:** To assess the technical feasibility and clinical applicability of SLNB with ICG under near-infrared (NIR) imaging guidance in patients with breast cancer.

**Materials and Methods:** This prospective single-center study included 30 patients who underwent ICG-guided SLNB between 2023 and 2025. In 24 patients, ICG mapping was combined with technetium-99m radioisotope navigation; in 6 patients, ICG guidance alone was used. The protocol comprised periareolar ICG injection, standardized imaging conditions, and fluorescence-index recording.

**Results:** The protocol was completed in all 30 patients. Fluorescent visualization of the lymphatic pathway and/or the sentinel lymph node (SLN) was achieved in every case, and no ICG-related adverse reactions were recorded. The mean fluorescence index was 213.0 *±* 24.7, the median was 206.0 [192.5–237.2], and the min–max was 180–255.

**Conclusion:** SLNB with ICG under NIR imaging guidance demonstrated technical feasibility in a prospective single-center cohort. Quantitative fluorescence-index recording may serve as a component of standardizing intraoperative fluorescence guidance.

## 1 INTRODUCTION

Breast cancer remains the most common malignant neoplasm in women and one of the leading causes of cancer-related mortality worldwide [1]. Staging of the regional lymphatic basin retains pivotal importance for selecting the extent of locoregional and systemic treatment; under contemporary conditions, however, it should be performed with the minimum necessary surgical trauma [2–4].

Sentinel lymph node biopsy (SLNB) is the principal tool for de-escalation of axillary surgery, providing morphological information on regional nodal status without primary removal of the entire axillary lymphatic basin [2]. In selected patients with a clinically negative axilla, randomized trials have shown that axillary surgery may even be safely omitted, reflecting the broader trend toward minimizing axillary intervention [5, 6]. Fluorescence lymphography with indocyanine green (ICG) is a promising method for intraoperative detection of the sentinel lymph node (SLN): it enables real-time visualization of lymphatic flow, does not require a blue dye, and may reduce the dependence of the procedure on radioisotope logistics. According to current evidence, ICG guidance is characterized by a high SLN detection rate and is comparable to conventional tracers across a variety of clinical scenarios in breast cancer [7–14].

At the same time, standardization of fluorescence-signal interpretation remains an unresolved problem, since the visual intensity of the signal depends on the lighting conditions, the distance to the camera, the depth of the node, the dye concentration, and the hardware parameters of the system. Therefore, for implementation of navigation using the MARS system in the near-infrared (NIR) range, not only visualization of the lymphatic pathway and the SLN is of fundamental importance, but also a quantitatively reproducible criterion of a sufficient fluorescence signal.

The aim of this study was to assess the technical feasibility and clinical applicability of SLNB with ICG under NIR imaging guidance in patients with breast cancer, and to characterize the distribution of the prospectively recorded fluorescence index under standardized conditions of intraoperative imaging.

## 2. MATERIALS AND METHODS

### 2.1. Study Design

The study was conducted as a prospective, single-center, non-randomized, observational study. The analysis included 30 patients with morphologically verified breast cancer who underwent SLNB with ICG fluorescence guidance.

### 2.2. Eligibility Criteria

The inclusion criteria were morphologically confirmed breast cancer; absence of distant metastases; indications for surgical treatment with assessment of regional nodal status; absence of contraindications to ICG; and signed informed consent.

The non-inclusion/exclusion criteria were generalization of the tumor process; severe comorbidity limiting surgical intervention; clinically significant coagulation disorders; contraindications to ICG; refusal to participate; and an incomplete set of intraoperative data.

### 2.3. Setting and Study Duration

The study was performed at the Department of Oncological Surgical Treatment Methods of University Clinical Hospital No. 1 of Sechenov University between 2023 and 2025. In 24 patients, ICG guidance was combined with technetium-99m radioisotope navigation; in 6 patients, ICG guidance alone was used. This distribution reflected the sequential introduction of the technology into clinical practice while adhering to a standardized protocol.

### 2.4. Intervention and Data Recording

For fluorescence guidance, ICG was used (Indocyanine Green lyophilizate, 25 mg, for the preparation of a solution for intravenous administration, single vial; Mirpharm, Russia) at a concentration of 2.5 mg/mL. After preparation of the surgical field, intradermal periareolar administration was performed, with 0.1 mL injected at each of four points corresponding to the conventional 12, 3, 6, and 9 o’clock positions; the total injected volume was 0.4 mL, and the total dose was 1.0 mg. After injection, gentle massage of the injection area was performed for 5 minutes.

Fluorescence-signal recording was performed using the MARS NIR fluorescence imaging system, software version 2.0.0.7 (Fig. 1). Within the system software, the fluorescence signal was quantified by placing a region of interest over the fluorescent sentinel node and over adjacent background tissue, from which the fluorescence index was derived (Fig. 2).

**Figure 1.**
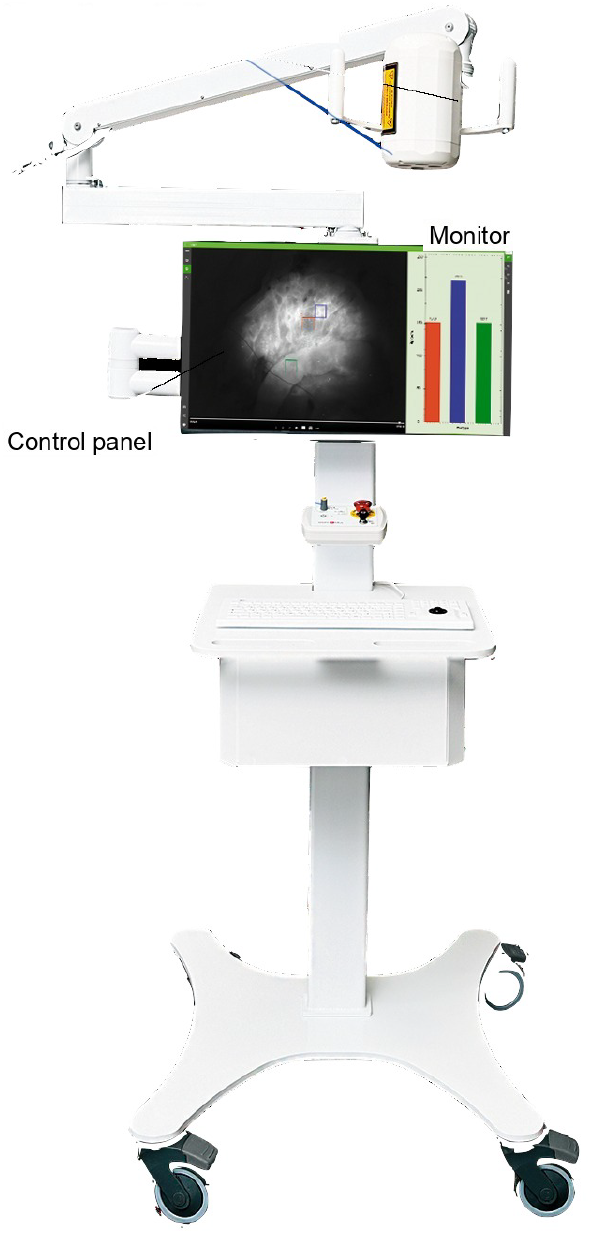
MARS near-infrared fluorescence imaging system used for intraoperative recording of the fluorescence signal.

**Figure 2.**
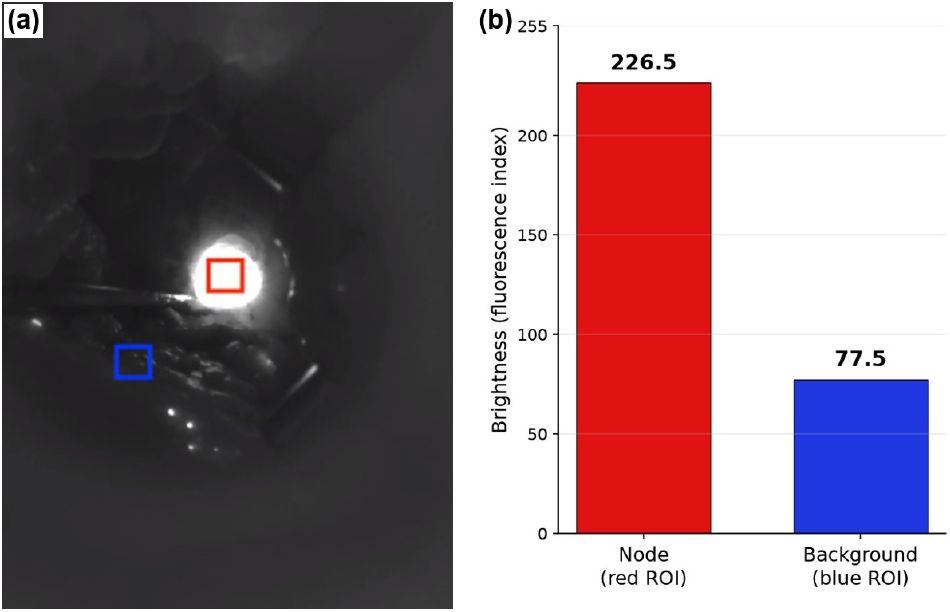
Quantification of the fluorescence signal in the MARS system software. (a) A region of interest is placed over the fluorescent sentinel lymph node (red box) and over adjacent background tissue (blue box). (b) The corresponding brightness readout for the node (red) and background (blue); the node value represents the fluorescence index.

Imaging was performed with minimized external lighting: the surgical lights were switched off or directed away from the operative field. The camera was positioned perpendicular to the skin or surgical wound at a distance of 30 cm. In real time, the lymphatic vessels were traced from the injection site to the first fluorescent SLN (Fig. 3).

**Figure 3.**
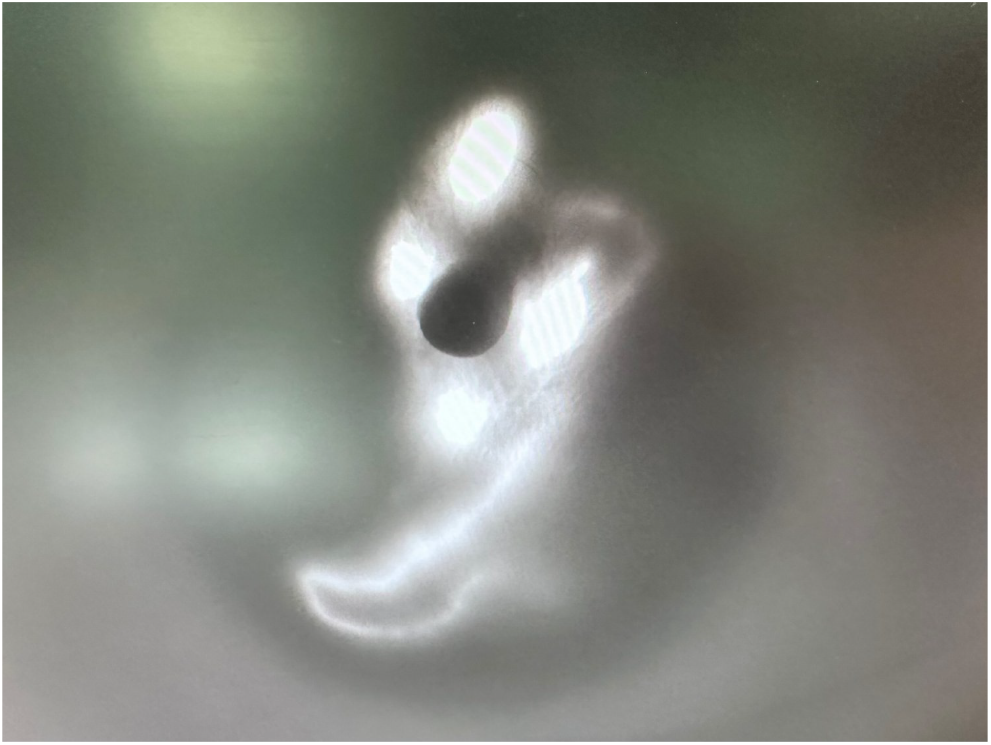
Intraoperative near-infrared fluorescence visualization of an indocyanine green–filled lymphatic channel draining toward the sentinel lymph node.

After removal of the first fluorescent node, the search was continued until the residual fluorescence in the axillary region had disappeared. The standardized parameters of fluorescence guidance are presented in Table 1.

**Table 1.**
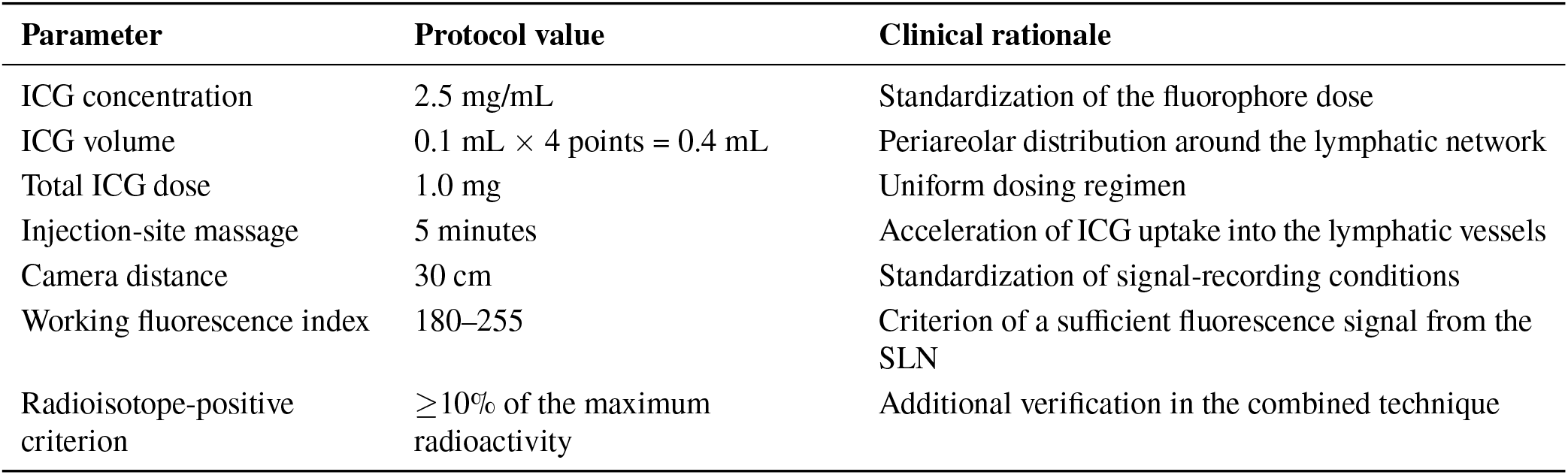
Standardized parameters of fluorescence guidance.

The working range of the fluorescence index (180–255) was established prior to statistical processing as a technical criterion of a sufficient fluorescence signal under standardized imaging conditions. The lower bound of 180 corresponded to the minimum signal level at which the lymphatic pathway and/or the SLN were confidently visualized by the operator. The value of 255 corresponded to the maximum value recorded in the analyzed series. This index was regarded as a technical visualization criterion rather than as a diagnostic criterion of metastatic lymph node involvement.

In the combined group, radioisotope navigation was performed using the radiopharmaceutical Nanotop labeled with technetium-99m. The radiopharmaceutical was administered 1–12 hours before surgery at a dose of 80–150 MBq, peritumorally, intradermally, or into the subareolar plexus. The route of administration, the interval before surgery, and the need for single-photon emission computed tomography combined with computed tomography (SPECT/CT) were determined in accordance with the local clinical protocol and the organizational conditions of the radioisotope stage. A node was considered radioisotope-positive when at least 10% of the maximum radioactivity level in the axillary region was recorded.

The fluorescence index was recorded intraoperatively for each patient during MARS/NIR mapping. The patient-specific data included the navigation modality, the parameters of the radioisotope stage when applied, the constant parameters of the fluorescence protocol, and the recorded fluorescence-index value. In all cases, the following were kept uniform: an ICG concentration of 2.5 mg/mL, a total injected volume of 0.4 mL, a total dose of 1.0 mg, an injection-site massage duration of 5 minutes, and a camera distance of 30 cm.

### 2.5. Study Endpoints

The primary endpoint was the technical feasibility of the protocol for SLNB with ICG under NIR imaging guidance. Technical feasibility was defined as the ability to perform fluorescence mapping under the specified conditions of drug dosing, standardized camera positioning, and fluorescence-signal recording.

The secondary descriptive endpoints were the rate of successful fluorescent visualization of the lymphatic pathway and/or the SLN; the occurrence of adverse reactions associated with ICG administration; the distribution of the fluorescence index; the proportion of values within the predefined working range of 180–255; and differences in the fluorescence index between protocol subgroups. For the between-group analysis, four subgroups were defined according to the route of radioisotope administration: ICG plus technetium-99m with subareolar administration; ICG plus technetium-99m with intradermal administration; ICG plus technetium-99m with peritumoral administration; and ICG guidance alone.

### 2.6. Statistical Analysis

Statistical analysis was performed using IBM SPSS Statistics version 26.0 (IBM Corp., Armonk, NY, USA). Quantitative data are presented as the mean *±* standard deviation (SD) and as the median [Q1–Q3]. Minimum and maximum values are presented as min–max. The normality of the fluorescence-index distribution was assessed using the Shapiro–Wilk test. For the proportion of observations within the predefined working range of the fluorescence index (180–255), an exact binomial 95% confidence interval (CI) was calculated. Between-group comparison of the fluorescence index across the protocol subgroups was performed using the Kruskal–Wallis test. This analysis was regarded as exploratory owing to the small number of observations and the non-randomized formation of the subgroups. Differences were considered statistically significant at *p <* 0.05. The *p* values are reported to three decimal places.

### 2.7. Ethics Review

The study protocol was approved by the Local Ethics Committee of I.M. Sechenov First Moscow State Medical University of the Ministry of Health of the Russian Federation (protocol No. 11-23 of 15 June 2023). Patient enrollment commenced after ethics approval. All patients provided written informed consent for surgical treatment, the use of ICG, and the use of de-identified clinical data for scientific purposes. The study was conducted in accordance with the principles of the Declaration of Helsinki of 1975 and its revised version of 2000, as well as with the ethical standards of the local ethics committee of the institution in which the work was carried out.

## 3. RESULTS

Thirty patients with breast cancer were included in the study. The median age was 62.0 years (min–max, 36–80 years). A clinically negative regional nodal status was determined in 22/30 (73.3%) patients. Combined navigation with ICG and technetium-99m was used in 24/30 (80.0%) patients, and ICG guidance without a radioisotope stage in 6/30 (20.0%) patients (Table 2).

**Table 2.** Clinical characteristics of the patients.

| Variable | Value |
| --- | --- |
| Number of patients | 30 |
| Age, years | Median 62.0;<br>min–max 36–80 |
| cN0 | 22 (73.3%) |
| cN1–cN3a | 8 (26.7%) |
| Luminal A | 14 (46.7%) |
| Luminal B | 13 (43.3%) |
| Other molecular subtypes | 3 (10.0%) |
| ICG + technetium-99m | 24 (80.0%) |
| ICG only | 6 (20.0%) |

Note. cN — clinical regional lymph node status.

The fluorescence guidance protocol was completed in all 30 patients. Fluorescent visualization of the lymphatic pathway and/or the SLN was achieved in every case. No adverse reactions associated with ICG administration were recorded. The intraoperative and ex-vivo steps of the ICG fluorescence workflow are illustrated in Fig. 4.

**Figure 4.**
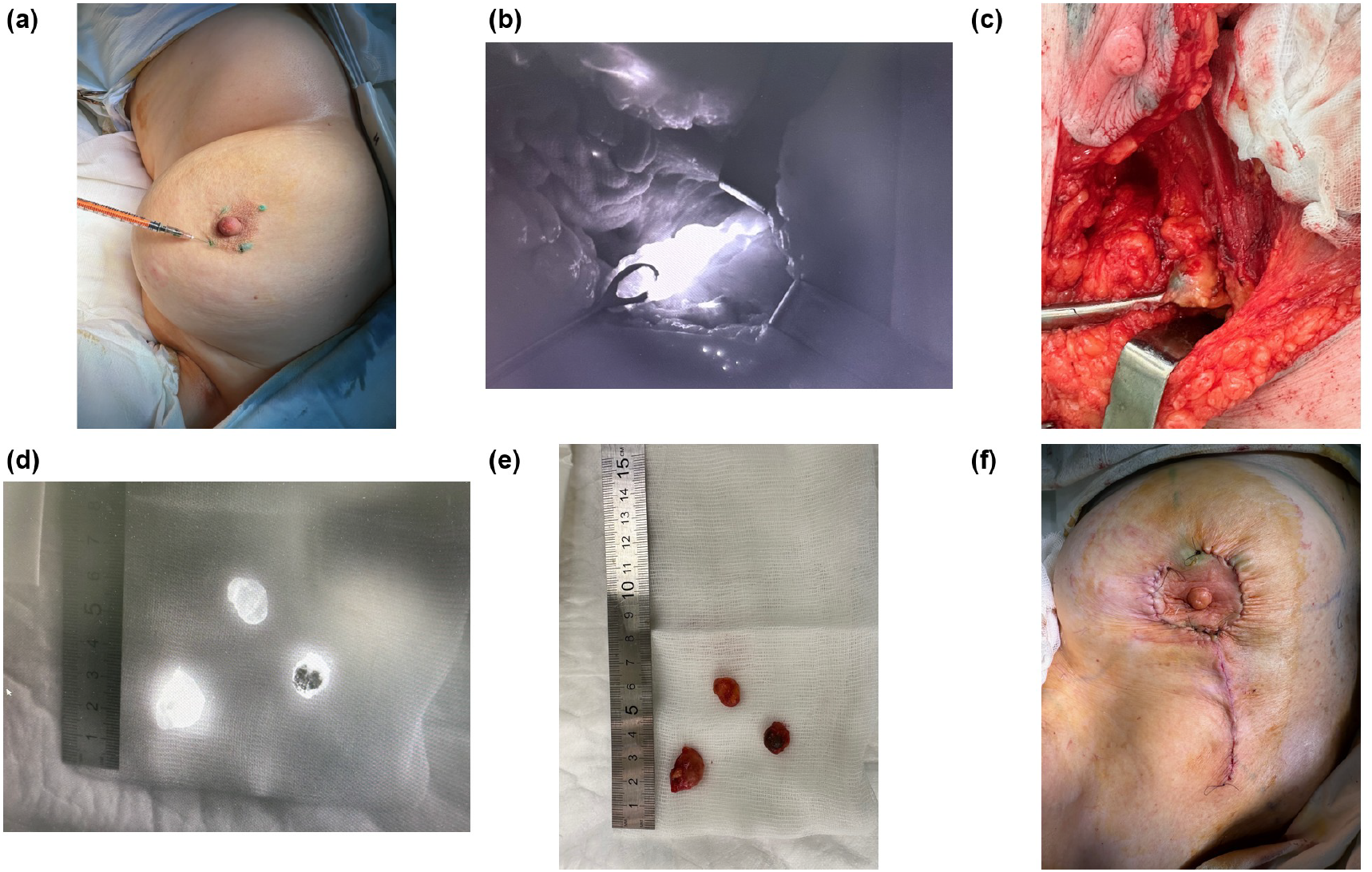
Intraoperative and ex-vivo indocyanine green (ICG) fluorescence workflow. (a) Periareolar intradermal injection of ICG; (b) intraoperative near-infrared visualization of the fluorescent sentinel lymph node during guided dissection; (c) gross appearance of the ICG-stained sentinel lymph node in the operative field; (d) excised sentinel lymph node under near-infrared fluorescence (with scale bar); (e) the same node in white light (with scale bar); (f) operative result after sentinel lymph node biopsy and lumpectomy.

The mean fluorescence index was 213.0 *±* 24.7 (95% CI, 203.7–222.2). The median was 206.0 [192.5–237.2], with a min–max of 180–255. All 30 values (100.0%) lay within the predefined working range of 180–255; the exact binomial 95% CI was 88.4–100.0% (Table 3).

**Table 3.** Fluorescence-index parameters.

| Parameter | Value |
| --- | --- |
| Mean ± SD | 213.0 ± 24.7 |
| 95% CI of the mean | 203.7–222.2 |
| Median [Q1–Q3] | 206.0<br>[192.5–237.2] |
| Min–max | 180–255 |
| Values within the working range | 30/30 (100.0%) |
| 95% CI for the proportion | 88.4–100.0% |
| Shapiro–Wilk test | $W = 0.904$;<br>$p = 0.011$ |
| Kruskal–Wallis test | $\chi^2 = 26.471$; df<br>$= 3$; $p < 0.001$ |

The distribution of the fluorescence index deviated from normal according to the Shapiro–Wilk test (*W* = 0.904; *p* = 0.011). In an exploratory comparison of the four protocol subgroups defined by the route of radioisotope administration, statistically significant differences in the fluorescence index were found (*χ*^2^ = 26.471; df = 3; *p* < 0.001); these subgroup differences are interpreted with caution in the Discussion. Patient-specific fluorescence-index values are presented in Supplementary Table 1.

## DISCUSSION

The present cohort demonstrates the technical feasibility of ICG fluorescence guidance with the MARS/NIR system in 30 patients with breast cancer. The principal focus of the study was standardization of the optical stage of SLNB: the ICG dose, the conditions of signal recording, the camera distance, and the quantitative range of the fluorescence index. The clinical profile of the cohort was consistent with the selective rationale of axillary staging, as most patients had a clinically negative regional nodal status. The findings are consistent with the current literature, which confirms the high detection rate of sentinel lymph nodes with ICG fluorescence guidance, its comparability with the radioisotope method, its advantage over blue dye, and the clinical applicability of dual tracing [7–15]. In this context, the value of the MARS/NIR approach lies in the possibility of quantitatively recording the fluorescence signal, which may reduce the subjectivity of intraoperative assessment and improve the comparability of results between operators and clinical series. The importance of standardizing acquisition conditions is underscored by earlier observations that the recorded fluorescence signal is sensitive to the ICG concentration, with excessively high concentrations causing fluorophore quenching [16]. At the ICG concentration used in the present protocol (2.5 mg/mL), reliable fluorescent visualization was nonetheless achieved in all patients with the MARS camera. Recent data likewise confirm that ICG can be used either as a standalone fluorescent tracer or as a component of dual navigation with technetium-99m, which is consistent with the mixed-protocol design of the present study [13–15]. Camera-based near-infrared platforms other than MARS have reported comparable performance; using a Karl Storz VITOM exoscope, ICG achieved a per-node detection rate of 97.7% versus 78.2% for technetium-99m and identified all node-positive patients, including one missed by the radioisotope [16].

### 4.1. Imaging Modalities for ICG-Guided Sentinel Node Detection

Fluorescence-guided SLN mapping with ICG in breast cancer was first described using a charge-coupled-device camera with 760-nm near-infrared excitation [17]; systems of this type were later commercialized, such as the Hamamatsu Photodynamic Eye. Since then, several imaging platforms have been applied to breast SLN detection. Open-field handheld near-infrared camera systems provide real-time transcutaneous and intraoperative visualization; in a randomized trial, ICG-based fluorescence combined with technetium-99m was assessed with and without patent blue dye, and fluorescence imaging outperformed the blue dye [18]. Exoscope-derived systems, such as the Karl Storz VITOM, deliver magnified extracorporeal fluorescence imaging of the operative field [16]. Hybrid-tracer strategies combine both modalities within a single injection: ICG-^99m^Tc-nanocolloid unites preoperative lymphoscintigraphy with intraoperative gamma-probe and near-infrared fluorescence guidance [19]. Across these platforms, meta-analytic data indicate that fluorescence imaging is at least comparable to radioisotope and blue-dye techniques for node identification [8]. A recurring limitation, however, is that most systems provide only qualitative visualization, leaving the judgment of signal adequacy operator-dependent. In this respect, integrated platforms such as MARS, which additionally return a quantitative fluorescence readout, offer a route to standardization; the fluorescence index evaluated in the present study is one such quantitative criterion.

### 4.2. Interpretation of Subgroup Differences

It should be emphasized that the present study was designed to evaluate the feasibility of ICG fluorescence imaging with the MARS/NIR camera rather than to compare tracer techniques. Technetium-99m, where used, served only as a concurrent standard-of-care confirmation of nodal localization and was not the object of investigation. Accordingly, the exploratory between-subgroup difference in the fluorescence index (Kruskal–Wallis *p <* 0.001) should be interpreted with caution. Because the optical acquisition parameters—ICG concentration, injected volume and dose, and camera distance—were held constant across all patients, this difference cannot be attributed to the fluorescence protocol itself. The subgroups were defined by the route and timing of radioisotope administration, which may co-vary with clinical factors such as nodal depth, body habitus, and lymphatic transit time that can plausibly influence the recorded signal; residual confounding and the small, non-randomized subgroup sizes preclude any causal interpretation. These subgroup observations are therefore hypothesis-generating only and warrant prospective validation with prespecified, blinded fluorescence-index measurement.

### 4.3. Study Limitations

The limitations of the study include the single-center design, the small sample size, the absence of randomization, the lack of independent blinded assessment of the fluorescence signal, and the mixed use of ICG both as a standalone tracer and in combination with technetium-99m. In addition, the present analysis did not assess the false-negative rate, long-term oncological outcomes, the inter-operator reproducibility of the measurements, or the relationship between the fluorescence index and the morphological status of each removed SLN. Accordingly, the present results should be regarded as confirmation of technical feasibility and as a basis for further validation of the standardized MARS/NIR protocol in larger prospective studies.

## 5. CONCLUSION

SLNB with ICG under NIR imaging guidance demonstrated technical feasibility within a standardized intraoperative protocol. In all cases, fluorescent visualization of the lymphatic pathway and/or the SLN was achieved, and the fluorescence-index values lay within the predefined working range. Quantitative recording of the fluorescence index may be considered a component of standardizing fluorescence guidance; further validation will require studies with a larger sample size and analysis of the concordance between fluorescent, radioisotope, and morphological data.

## Supporting information

Supplementary Table 1. Patient-level MARS fluorescence-index values.

## Data Availability

All data produced in the present study are available upon reasonable request to the authors.

## Author Contributions

F. Kurdi, Y. Kurdi, and I. V. Reshetov — study concept and design; F. Kurdi, Y. Kurdi, M. Kurdi, A. A. Shiryaev, N. S. Sukortseva, and T. N. Pisareva — collection and systematization of the clinical data; F. Kurdi, Y. Kurdi, A. L. Istranov, and I. V. Reshetov — surgical and clinical support; F. Kurdi and Y. Kurdi — statistical analysis; Y. Kurdi and F. Kurdi — manuscript preparation. All authors participated in editing and read and approved the final version of the manuscript.

## Ethical Statement

The study protocol was approved by the Local Ethics Committee of I.M. Sechenov First Moscow State Medical University of the Ministry of Health of the Russian Federation (protocol No. 11-23 of 15 June 2023). Patient enrollment commenced after ethics approval. All patients provided written informed consent for surgical treatment, the use of ICG, and the use of de-identified clinical data for scientific purposes. The study was conducted in accordance with the principles of the Declaration of Helsinki of 1975 and its revised version of 2000. No animal experiments were performed.

