## Supplementary Table 1. Patient-level MARS fluorescence-index values. for "Indocyanine Green Fluorescence-Guided Sentinel Lymph Node Biopsy in Breast Cancer Using the MARS Near-Infrared Imaging System: A Prospective Single-Center Feasibility Study"

In all patients the optical acquisition parameters were held constant (indocyanine green [ICG] 2.5 mg/mL; total injected volume 0.4 mL; total dose 1.0 mg; camera distance 30 cm). The working fluorescence-index range was predefined as 180–255. Technetium-99m ($^{99m}$Tc) was used only for concurrent standard-of-care nodal confirmation and was not the object of investigation.

| No. | Method | $^{99m}$Tc: route; time; dose | ICG/NIR constants | Index | Interpretation |
|---|---|---|---|---|---|
| 1 | ICG + $^{99m}$Tc | Subareolar; 1 h; 80 MBq | 2.5 mg/mL; 0.4 mL; 30 cm | 255 | High signal |
| 2 | ICG + $^{99m}$Tc | Subareolar; 1 h; 80 MBq | 2.5 mg/mL; 0.4 mL; 30 cm | 248 | High signal |
| 3 | ICG + $^{99m}$Tc | Subareolar; 1 h; 80 MBq | 2.5 mg/mL; 0.4 mL; 30 cm | 244 | High signal |
| 4 | ICG + $^{99m}$Tc | Subareolar; 1 h; 80 MBq | 2.5 mg/mL; 0.4 mL; 30 cm | 252 | High signal |
| 5 | ICG + $^{99m}$Tc | Subareolar; 1 h; 80 MBq | 2.5 mg/mL; 0.4 mL; 30 cm | 239 | High signal |
| 6 | ICG + $^{99m}$Tc | Subareolar; 1 h; 80 MBq | 2.5 mg/mL; 0.4 mL; 30 cm | 247 | High signal |
| 7 | ICG + $^{99m}$Tc | Subareolar; 1 h; 80 MBq | 2.5 mg/mL; 0.4 mL; 30 cm | 251 | High signal |
| 8 | ICG + $^{99m}$Tc | Subareolar; 1 h; 80 MBq | 2.5 mg/mL; 0.4 mL; 30 cm | 243 | High signal |
| 9 | ICG + $^{99m}$Tc | Intradermal; 6 h; 100 MBq | 2.5 mg/mL; 0.4 mL; 30 cm | 190 | Threshold-sufficient |
| 10 | ICG + $^{99m}$Tc | Intradermal; 6 h; 100 MBq | 2.5 mg/mL; 0.4 mL; 30 cm | 198 | Threshold-sufficient |
| 11 | ICG + $^{99m}$Tc | Intradermal; 6 h; 100 MBq | 2.5 mg/mL; 0.4 mL; 30 cm | 205 | Stable signal |
| 12 | ICG + $^{99m}$Tc | Intradermal; 6 h; 100 MBq | 2.5 mg/mL; 0.4 mL; 30 cm | 194 | Threshold-sufficient |
| 13 | ICG + $^{99m}$Tc | Intradermal; 6 h; 100 MBq | 2.5 mg/mL; 0.4 mL; 30 cm | 201 | Stable signal |
| 14 | ICG + $^{99m}$Tc | Intradermal; 6 h; 100 MBq | 2.5 mg/mL; 0.4 mL; 30 cm | 207 | Stable signal |
| 15 | ICG + $^{99m}$Tc | Intradermal; 6 h; 100 MBq | 2.5 mg/mL; 0.4 mL; 30 cm | 196 | Threshold-sufficient |
| 16 | ICG + $^{99m}$Tc | Intradermal; 6 h; 100 MBq | 2.5 mg/mL; 0.4 mL; 30 cm | 202 | Stable signal |
| 17 | ICG + $^{99m}$Tc | Peritumoral; 12 h; 150 MBq | 2.5 mg/mL; 0.4 mL; 30 cm | 180 | Threshold-sufficient |
| 18 | ICG + $^{99m}$Tc | Peritumoral; 12 h; 150 MBq | 2.5 mg/mL; 0.4 mL; 30 cm | 184 | Threshold-sufficient |
| 19 | ICG + $^{99m}$Tc | Peritumoral; 12 h; 150 MBq | 2.5 mg/mL; 0.4 mL; 30 cm | 188 | Threshold-sufficient |
| 20 | ICG + $^{99m}$Tc | Peritumoral; 12 h; 150 MBq | 2.5 mg/mL; 0.4 mL; 30 cm | 192 | Threshold-sufficient |
| 21 | ICG + $^{99m}$Tc | Peritumoral; 12 h; 150 MBq | 2.5 mg/mL; 0.4 mL; 30 cm | 186 | Threshold-sufficient |
| 22 | ICG + $^{99m}$Tc | Peritumoral; 12 h; 150 MBq | 2.5 mg/mL; 0.4 mL; 30 cm | 190 | Threshold-sufficient |
| 23 | ICG + $^{99m}$Tc | Peritumoral; 12 h; 150 MBq | 2.5 mg/mL; 0.4 mL; 30 cm | 181 | Threshold-sufficient |
| 24 | ICG + $^{99m}$Tc | Peritumoral; 12 h; 150 MBq | 2.5 mg/mL; 0.4 mL; 30 cm | 195 | Threshold-sufficient |
| 25 | ICG only | Not applied | 2.5 mg/mL; 0.4 mL; 30 cm | 222 | Stable signal |
| 26 | ICG only | Not applied | 2.5 mg/mL; 0.4 mL; 30 cm | 214 | Stable signal |
| 27 | ICG only | Not applied | 2.5 mg/mL; 0.4 mL; 30 cm | 226 | Stable signal |
| 28 | ICG only | Not applied | 2.5 mg/mL; 0.4 mL; 30 cm | 218 | Stable signal |
| 29 | ICG only | Not applied | 2.5 mg/mL; 0.4 mL; 30 cm | 209 | Stable signal |
| 30 | ICG only | Not applied | 2.5 mg/mL; 0.4 mL; 30 cm | 232 | High signal |

*Notes.* Interpretation categories are descriptive labels for the recorded fluorescence-index level: threshold-sufficient, 180–199; stable signal, 200–229; high signal, $\geq$230. All 30/30 (100.0%) values fell within the predefined working range of 180–255. Subgroups are defined by the route of $^{99m}$Tc administration and are provided for the exploratory analysis reported in the main text; because all optical acquisition parameters were held constant, between-subgroup differences are hypothesis-generating only. Aggregate statistics (mean 213.0 $\pm$ 24.7; median 206.0 [192.5–237.2]; Shapiro–Wilk $W = 0.904$, $p = 0.011$; Kruskal–Wallis $\chi^2 = 26.471$, df = 3, $p < 0.001$) are reported in Table 3 of the main article.
